# Prevalence and risk factors of self-reported audiovestibular symptoms in a population-based sample from rural northeastern Germany

**DOI:** 10.1101/2023.08.17.23293690

**Authors:** Friedrich Ihler, Tina Brzoska, Reyhan Altindal, Oliver Dziemba, Henry Völzke, Chia-Jung Busch, Till Ittermann

## Abstract

**Objectives:** The senses of hearing and balance are linked by a close anatomical and physiological relationship. A further pathophysiological interaction is supposed but the detailed mechanism and direction remains elusive. Further insight is required into the prevalence of the audiovestibular symptoms hearing loss, tinnitus and dizziness as only scarce information on the combined occurrence is available so far. Therefore, this was assessed in a population-based sample. Based on this, we studied the influence of risk factors from lifestyle habits as well as cardiovascular and metabolic conditions on the development of audiovestibular symptoms alone and in combination.

**Design:** This analysis evaluated the prevalence of self-reported hearing loss, tinnitus and dizziness in two separate population-based samples from West Pomerania, a rural region of north-eastern Germany. Datasets from 8134 individuals aged 20 to 79 years were available from the baseline investigations of the cohorts START and TREND of the Study of Health in Pomerania (SHIP). Audiovestibular symptoms were assessed by structured questionnaires. The cohorts were comprehensively characterized regarding modifiable lifestyle factors as well as cardiovascular and metabolic disorders, allowing the assessment of the role of those influencing factors.

**Results:** Audiovestibular symptoms were prevalent and overlapping in the investigated population. 2350 individuals (28.9%) reported at least one, 648 (8.1%) two and 111 (1.4%) all three audiovestibular symptoms. Thereby, we observed a weighted prevalence of 14.2% for hearing loss, 9.7% for tinnitus and 13.5% for dizziness in the population. The prevalences increased with age and differed among the sexes. The prevalence of hearing loss as well as tinnitus increased between the two cohorts. A moderate positive correlation was found between hearing loss and tinnitus (phi-coefficient 0.318). In multivariable regression analyses, only smoking was significantly associated with all three symptoms. Less education and several cardiovascular risk factors contributed to both hearing loss and dizziness.

**Conclusions:** Audiovestibular symptoms are highly prevalent in the general population and the occurrence overlaps. A considerable but complex influence of risk factors points towards a relation with neuronal as well as cardiovascular disease processes. Future studies should identify subgroups that are particularly at risk. Additionally, to clarify the underlying mechanisms the interaction between the senses of hearing and balance as well as the mode of action of the risk factors should be evaluated in more detail in the future.

## Introduction

The inner ear contains the sensory organ of the auditory system as well as the vestibular labyrinth. While the hearing sense detects sound, the peripheral vestibular organ is stimulated by either linear or angular acceleration. The peripheral vestibular organ is thereby one of the main contributing modalities of the more complex vestibular system which in turn constitutes the sense of balance. Hearing loss and tinnitus are symptoms for dysfunction of the auditory system while dizziness is a general term for the subjective experience during balance disturbance. Those audiovestibular symptoms are each considerably prevalent when viewed individually, with hearing loss affecting up to 5% (Sheffield & Smith 2019), tinnitus up to 42.7% (McCormack et al. 2016) an dizziness up to 30% (Murdin & Schilder 2015) of investigated populations.

Heterogenous but rare disorders of the cerebellopontine angle or the temporal bone are known to lead to various combinations of the three symptoms of audiovestibular dysfuncion, including vestibular schwannoma (Goldbrunner et al. 2020), Menière’s disease (Lopez-Escamez et al. 2015) or superior semicircular canal dehiscence syndrome (Ward et al. 2021). Beyond this, ample evidence points towards a relevant co-prevalence of audiovestibular symptoms. Thereby, hearing loss is seen as a major risk factor for tinnitus (Baguley et al. 2013; Biswas & Hall 2021) and accordingly, the rehabilitation of hearing loss by sound-amplifying hearing aids can provide relieve from tinnitus symptoms (Jacquemin et al.; Baguley et al. 2013). Tinnitus may even be an early sign of cochlear dysfunction that might initially be out of the detection range of common auditory tests (Jafari et al. 2022). Likewise, during long-term follow-up, initially normal-hearing individuals with tinnitus progress to hearing loss in a large share of cases (Kehrle et al. 2022). Regarding hearing and balance, it is assumed that the hearing system recognizes spatial information (Zhong & Yost 2013; Rumalla et al. 2015; Vitkovic et al. 2016). Vice versa, there is evidence that auditory cues are processed during the perception of balance (Palm et al. 2009; Vitkovic et al. 2016) and are therefore likely to play a role in postural stability (Lubetzky et al. 2020). More specifically, hearing loss may have a detrimental effect on balance control (Carpenter & Campos 2020) and hearing rehabilitation has a beneficial effect on posture and gait (Rumalla et al. 2015; Vitkovic et al. 2016; Negahban et al. 2017; Louza et al. 2019). Finally, anxiety as well as phobic, somatoform and affective disorders are prevalent in individuals with organic or non-organic disorders of the vestibular system (Lahmann et al. 2015) as well as in those suffering from tinnitus (Baguley et al. 2013; Biswas & Hall 2021).

The assumption of a tight interaction between hearing and balance is substantiated further by various shared risk factors, with age being a major determinant. Inflammatory aging processes, age-related oxidative stress and hereditary factors are strongly supposed to contribute to a steeply increasing prevalence of auditory and vestibular dysfunction with advancing age (Paplou et al. 2021). However, while presbycusis is a complex but commonly accepted concept (Yamasoba et al. 2013; Eckert et al. 2021), a comparable interaction between age and vestibular dysfunction is less well established so far (Schubert et al. 2022). Also, a strong association is assumed between hearing loss and cognitive decline (Uchida et al. 2019). The complex process of neurodegeneration is viewed as a common underlying pathomechanism of central and peripheral auditory dysfunction as well as simultaneously being signified early by hearing difficulties and reinforced by sensory deprivation (Johnson et al. 2021).

Multiple modifiable lifestyle factors have been associated with the development of auditory or vestibular dysfunction but traditionally hearing loss, tinnitus and dizziness were considered separately. For hearing loss alone, low dietary quality (Le Prell et al. 2011; Spankovich & Le Prell 2013; Curhan et al. 2014; Jung et al. 2019; Dillard et al. 2022) and physical inactivity (Curhan et al. 2013; Tsimpida et al. 2019; Tseng et al. 2022) have been identified as specific risk factors. Alcohol consumption (Tsimpida et al. 2019; Biswas & Hall 2021) and obesity (Curhan et al. 2013; Hu et al. 2020; Biswas et al. 2021; Biswas & Hall 2021; Mick et al. 2023) might be contributing both to hearing loss (Curhan et al. 2013; Tsimpida et al. 2019; Hu et al. 2020; Mick et al. 2023) and tinnitus (Biswas et al. 2021; Biswas & Hall 2021). It is remarkable that only smoking seems to be a shared risk factor of all three audiovestibular symptoms hearing loss (Cruickshanks et al. 1998; Zhan et al. 2011; Lin et al. 2013; Tsimpida et al. 2019; Engdahl, Stigum, & Aarhus 2021; Tseng et al. 2022; Mick et al. 2023), tinnitus (Biswas et al. 2021; Biswas & Hall 2021; Goderie et al. 2022) and dizziness (Neuhauser et al. 2005; Agrawal et al. 2009) and might therefore be of particular importance.

As smoking is a major cardiovascular risk factor, it is important to consider that a strong association between auditory or balance dysfunction as well as cardiovascular disorders have been described. This is exemplified by stroke, as individuals with sensorineural hearing loss face an increased risk of such a cerebrovascular event (Khosravipour & Rajati 2021). This risk is even higher after the occurrence of sudden sensorineural hearing loss (Khosravipour & Rajati 2021; Lammers et al. 2021). In a cohort study, the presence of a cardiovascular risk profile was associated with the presence of hearing loss at baseline. Moreover, normal-hearing individuals at baseline had an increased risk for developing hearing loss during a 10-year follow-up period (Tseng et al. 2022). Furthermore, dizziness is one of the most common unspecific symptoms for stroke (Jones et al. 2021).

So far, prevalence and risk factors have been studied separately for individual audiovestibular symptoms. Given the close relationship between the senses of hearing and balance, the observation of an association of several risk factors with more than one audiovestibular symptom and a considerable mechanistic overlap between the risk factors, a more comprehensive analysis is warranted. Therefore, this study assessed the prevalence of self-reported hearing loss, dizziness and tinnitus in the baseline examinations of the SHIP-START and SHIP-TREND cohorts and the interaction of those three symptoms. Beyond this, it was analyzed whether their occurrence is influenced by age, sex, education, cardiovascular or metabolic risk factors as well as lifestyle habits.

## Methods

### Compliance with ethical and quality standards

The studies were conducted in accordance with the Declaration of Helsinki. The study protocols were approved by the responsible local ethics committee at the University Medicine Greifswald, Germany (approvals from July 31, 1995 (SHIP-START) and June 06, 2008 (BB 39/08, SHIP-TREND)). All participants gave written informed consent.

To comply with the aims of the EQUATOR (Enhancing the QUAlity and Transparency Of health Research) Network (http://www.equator-network.org/home/), this manuscript was prepared according to the Strengthening the Reporting of Observational Studies in Epidemiology (STROBE) statement (Elm et al. 2008). The completed checklist on recommended items is given as Supplemental Table 1.

### Study design and setting

The study population derived from the baseline examinations of two independent population-based samples of the Study of Health in Pomerania (SHIP). Adults aged 20–79 at the time of sampling were drawn from mainland West Pomerania, a rural area in northeastern Germany. The study region with approximately 220 thousand adult inhabitants comprises parts of the administrative districts of Vorpommern-Rügen and Vorpommern-Greifswald within the of the Federal State of Mecklenburg-West Pomerania. A time lag due to the process of sampling as well invitation, scheduling and conduction of investigations led to a maximum age of 81 at baseline.

The first cohort, SHIP-START, was recruited and underwent baseline investigations (designation for baseline SHIP-START-0) between 1997 and 2001, the second, SHIP-TREND, between 2008 and 2012 (SHIP-TREND-0). In brief, SHIP-START-0 was a stratified cluster-random sample of 7008 individuals; a net sample (without migrated or deceased persons) of 6265 eligible individuals; and 4308 (thereof 2192 women) who participated (response 68.8%). For SHIP-TREND-0, a stratified sample of 10000 individuals (net sample size of 8826) was drawn and 4420 (thereof 2275 women) participated (response 50.1%) (Schmidt et al. 2011).

SHIP is not focused on a specific disease or organ system, but instead on health and disease in general, resulting in self-report paper–pencil questionnaires, computer-assisted personal interviews as well as a comprehensive examination program in each examination wave. The study design and protocols have been described in detail elsewhere (Völzke et al. 2011; Völzke et al. 2022). Here, the data from the baseline examinations was studied, thereby representing a cross-sectional analysis.

### Variables

#### Outcome variables

Primary outcome measures were the self-reported occurrence of the audiovestibular symptoms hearing loss, tinnitus and dizziness in the study population. Grade 2 or 3 was regarded as severity of clinical relevance for consecutive analyses. The exact wording in German with English translation is given in Supplemental Table 2.

#### Exposure variables

The age at the day of baseline investigation was used for further analyses. Sex was documented in binary categories. Education was assessed during an interview and graded along the German educational framework.

Behavioral risk factors included smoking status (never/former, or current smoker). Pack years were calculated as average number of cigarettes smoked per day multiplied by the number of years smoking divided by 20. Alcohol consumption was assessed in grams ethanol per day derived from a quantity– frequency questionnaire (Baumeister et al. 2005). Physical inactivity was defined as 1 h leisure time physical activity or less per week.

Body mass index (BMI) and waist-to-hip-ratio were obtained via standardized measurement of body weight and height as well as waist and hip circumference and subsequent calculation. Diabetes mellitus was defined as insulin resistance >8.0 mmol/L non-fasting glucose or reported diabetes or reported diabetes treatment and/or dietary treatment. Arterial hypertension was defined as increased systolic or diastolic blood pressure >140/90 mmHg or reported hypertension/reported antihypertensive medication.

Metabolic syndrome was defined using a modification of the approach suggested by Alberti and coworkers as a combination of three out of five parameters: present diabetes mellitus as defined above; abdominal obesity: waist circumference > 94 cm (male) or > 80 cm (female); low HDL-cholesterol: < 1.03 mmol/L (male) or < 1.3 mmol/L (female); high triglycerides: > 2.3 mmol/L non-fasting triglycerides or lipid-lowering medication; present hypertension as defined above (Alberti et al. 2009; Schipf et al. 2010; Buchmann et al. 2022).

For laboratory parameters, random blood samples were collected without stasis from the cubital vein following a standardized protocol, refrigerated to 4-8°C and shipped on refrigerant packing within 4 to a maximum of 6h to the laboratory. Serum levels of glucose, total cholesterol, HDL cholesterol, and triglycerides were measured using the Dimension Vista 500 analytical system (Siemens Healthcare Diagnostics, Eschborn, Germany). Dyslipidemia was defined as intake of lipid-lowering medication (ATC C10) or increased levels of total cholesterol (≥6.2 mmol/L), total cholesterol-HDL-cholesterol-ratio (≥5.0) or LDL-cholesterol (≥4.1 mmol/L)

### Participants

In total, 8727 persons underwent baseline investigations in the cohorts SHIP-START (n= 4308) and SHIP-TREND (n= 4420). One participant from SHIP-START withdrew consent. The outcome variables were not completed by 593 participants (SHIP-START n= 93, SHIP-TREND n= 500). Therefore, datasets with the primary variables were available for this analysis from 8134 individuals (93.2%). Details on the study population are given in Supplemental Table 3.

### Analysis and reporting of data

Weighting was used to adjust for bias due to differences in responses, probabilities of selection, and discrepancies between data from official statistics and our samples with regard to demographic and geographical distributions (Schmidt et al. 2011). As different ways were used to obtain the samples in SHIP-START-0 and SHIP-TREND-0, all data except those in Table 1 were standardized using post stratification weighting. The factors included in the post stratification weighting were age, sex, and the data of the local registration office. Based on the data of the non-responder survey, the probability of participation in SHIP-TREND-0 was estimated by means of logistic regression. The resulting inverse probability weights were multiplied by the post stratification weights in SHIP-TREND-0.

**Table 1:** Characteristics of the study population stratified by grade 2 symptoms.

|  | No grade 2 symptom | At least one grade 2 symptom | p* |
| --- | --- | --- | --- |
| **Age; years** | 47 (35; 60) | 60 (47; 70) | <0.001 |
| **Females** | 50.9% | 50.5% | 0.735 |
| **Years of schooling** |  |  | <0.001 |
| < 10 years | 25.1% | 44.3% |  |
| 10 years | 51.0% | 40.4% |  |
| > 10 years | 23.9% | 15.3% |  |
| **Smoking status** |  |  | <0.001 |
| Never | 35.8% | 36.5% |  |
| Former | 32.8% | 40.9% |  |
| Current | 31.4% | 22.6% |  |
| **Alcohol consumption; g/day** | 5.05 (1.30; 13.4) | 3.59 (0.65; 11.5) | <0.001 |
| **Body mass index; kg/m<sup>2</sup>** | 26.8 (23.7; 30.2) | 27.8 (24.7; 31.0) | <0.001 |
| **Waist circumference; cm** | 89 (79; 99) | 93 (82; 102) | <0.001 |
| **Myocardial infarction** | 2.1% | 5.6% | <0.001 |
| **Stroke** | 1.4% | 4.2% | <0.001 |
| **Glucose; mmol/L** | 5.3 (4.9; 5.8) | 5.4 (5.0; 6.1) | <0.001 |
| **HbA1c; %** | 5.2 (4.8; 5.6) | 5.4 (5.0; 5.9) | <0.001 |
| **Type 2 diabetes** | 6.9% | 13.6% | <0.001 |
| **Total cholesterol; mmol/L** | 5.3 (4.6; 6.1) | 5.4 (4.7; 6.2) | 0.024 |
| **HDL-cholesterol; mmol/L** | 1.33 (1.09; 1.60) | 1.30 (1.06; 1.58) | 0.009 |
| **LDL-cholesterol; mmol/L** | 3.3 (2.6; 3.9) | 3.3 (2.7; 4.0) | 0.102 |
| **Triglycerides; mmol/L** | 1.40 (0.97; 2.05) | 1.54 (1.10; 2.24) | <0.001 |
| **Dyslipidemia** | 17.6% | 30.5% | <0.001 |
| **Arterial hypertension** | 44.8% | 60.0% | <0.001 |

Continuous data are expressed as median, 25<sup>th</sup> and 75<sup>th</sup> percentile; categorical data as percentage.
\*: Mann-Whitney-U test (continuous data) or $\chi^2$ -test (categorical data).

Prevalence of audiovestibular symptoms were reported stratified by sex and age groups. Correlations between the audiovestibular symptoms hearing loss, tinnitus, and dizziness were calculated by phi-coefficients. Stratified by grade 2 symptoms, characteristics of the study population were reported as median, 25th and 75th percentile for continuous data and as percentage for categorical data. Group comparisons were conducted by Mann-Whitney U-tests for continuous data and by χ2-tests for categorical data. Associations of behavioral and metabolic markers with audiovestibular symptoms were calculated by logistic regression models adjusted for age, sex, and study cohort. To make the odds ratios comparable, continuous exposure variables were standardized before usage in the regression models. In all analyses a p<0.05 was considered as statistically significant. Analyses were done with Stata 17.0 (Stata Corporation; College Station, USA).

## Results

In our study population we observed a relevant severity (grade 2 or 3) of audiovestibular symptoms. Out of 8134 individuals, 1190 (14.6%) suffered from hearing loss, 815 (10.0%) from tinnitus and 1114 (13.7%) from dizziness. There was a considerable overlap between symptoms, with 2350 (28.9%) reporting at least one, 648 (8.1%) two and 111 (1.4%) all three audiovestibular symptoms. Of these, 28.0% of cases with symptoms (648/2350) reported more than one. A breakdown by individual symptoms is given in Figure 1A. Considering possible sampling bias, we derived a weighted prevalence of 14.2% for hearing loss, 9.7% for tinnitus and 13.5% for dizziness in the population. Of note, a weighted prevalence of 28.0% for any audiovestibular symptom of relevant grade was considerably high. Occurrence of symptoms varied depending on sex, with hearing loss being more prevalent in males and dizziness in females (Figure 1B).

**Figure 1:**
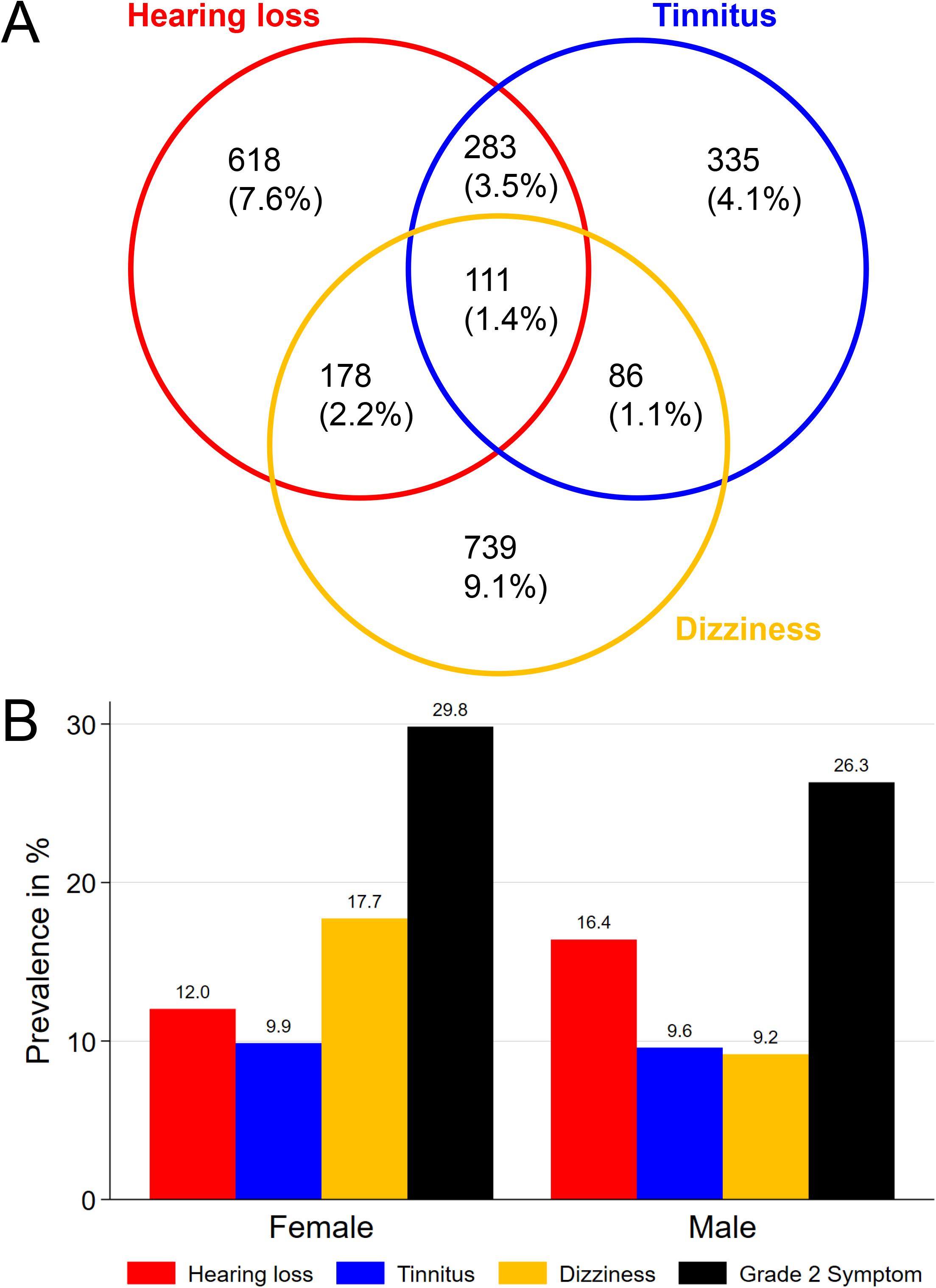
Prevalence of audiovestibular symptoms. A: individual and multiple symptoms (absolute numbers and share from total study population, n = 8134), B: weighted prevalence, individual and any grade-2-symptom.

Interestingly, an increase of the prevalence of audiovestibular symptoms was noted when the two cohorts SHIP-START and SHIP-TREND were compared. Between the cohorts, the prevalence for hearing loss increased significantly from 14.1% (95% CI 12.5; 15.6) to 17.9% (95% CI 16.1; 19,7) in males (p= 0.001) and from 9.8% (95% CI 8.5; 11.1) to 13.5% (95% CI 11.7; 15.2) in females (p= 0.001). Likewise, the prevalence for tinnitus increased significantly from 8.3% (95% CI 7.1; 9.5) to 10.4% (95% CI 9.0; 11.8) in males (p= 0.022) and from 7.2% (95% CI 6.1; 8.4) to 11.6% (95% CI 10.0; 13.2) in females (p< 0.001). In contrast, the prevalence of dizziness showed an increase from 7.8% (95% CI 6.6; 9.1) to 10.0% (95 % CI 8.6; 11.5) in males (p= 0.023), but decreased from 19.8% (95 % CI 18.0; 21.6) to 16.4% (95 % CI 14.5, 18.2) in females (p= 0.011). Thereby, the prevalence of at least one grade 2 symptom increased significantly from 23.1% (95 % CI 21.1; 25.0) to 28.4% (95 % CI 26.3; 30.5) in males (p< 0.001). This has brought the prevalence closer towards the level of females, where it has not changed significantly between cohorts. The reason for changes within little more than a decade is not fully clear.

There was also a trend towards symptoms influencing each other. The correlation for the co-occurrence of relevant symptoms (phi-coefficient) was 0.318 for hearing loss and tinnitus, 0.127 for hearing loss and dizziness as well as 0.102 for tinnitus and dizziness. This represents a moderate positive relationship between hearing loss and tinnitus that is comparatively stronger than the two other pairings.

We stratified the study cohort according to the absence or presence of grade 2 symptoms. While sex was evenly distributed here, individuals with grade 2 symptoms were significantly older. A wide range of lifestyle habits as well as metabolic or cardiovascular risk factors and disorders were significantly associated with the presence of symptoms. An overview is given in Table 1. Furthermore, the presence of relevant individual and multiple audiovestibular symptoms increased with age. In 2614 individuals with an age of 60 or older, 1170 (44.8%) reported at least one, 391 (15.0%) two and 69 (2.6%) all three symptoms. This was even more so in 1097 individuals at the age of 70 or older with 579 (52.8%) reporting at least one, 218 (19.9%) two and 38 (3.5%) all three. This trend was also observed when weighted prevalence was broken down by age and sex. In the oldest age group of 70 to 81 years at the day of examinations, the majority of individuals reported at least one grade 2 symptom in both sexes. Dizziness was leading in females until being overtaken by hearing loss in the oldest age group (Figure 2A). In contrast, the dominant symptom in males was hearing loss from the age group of 30 to 39 on, increasing from a weighted prevalence of 6.2% to 41.7% in the oldest group (Figure 2B).

**Figure 2:**
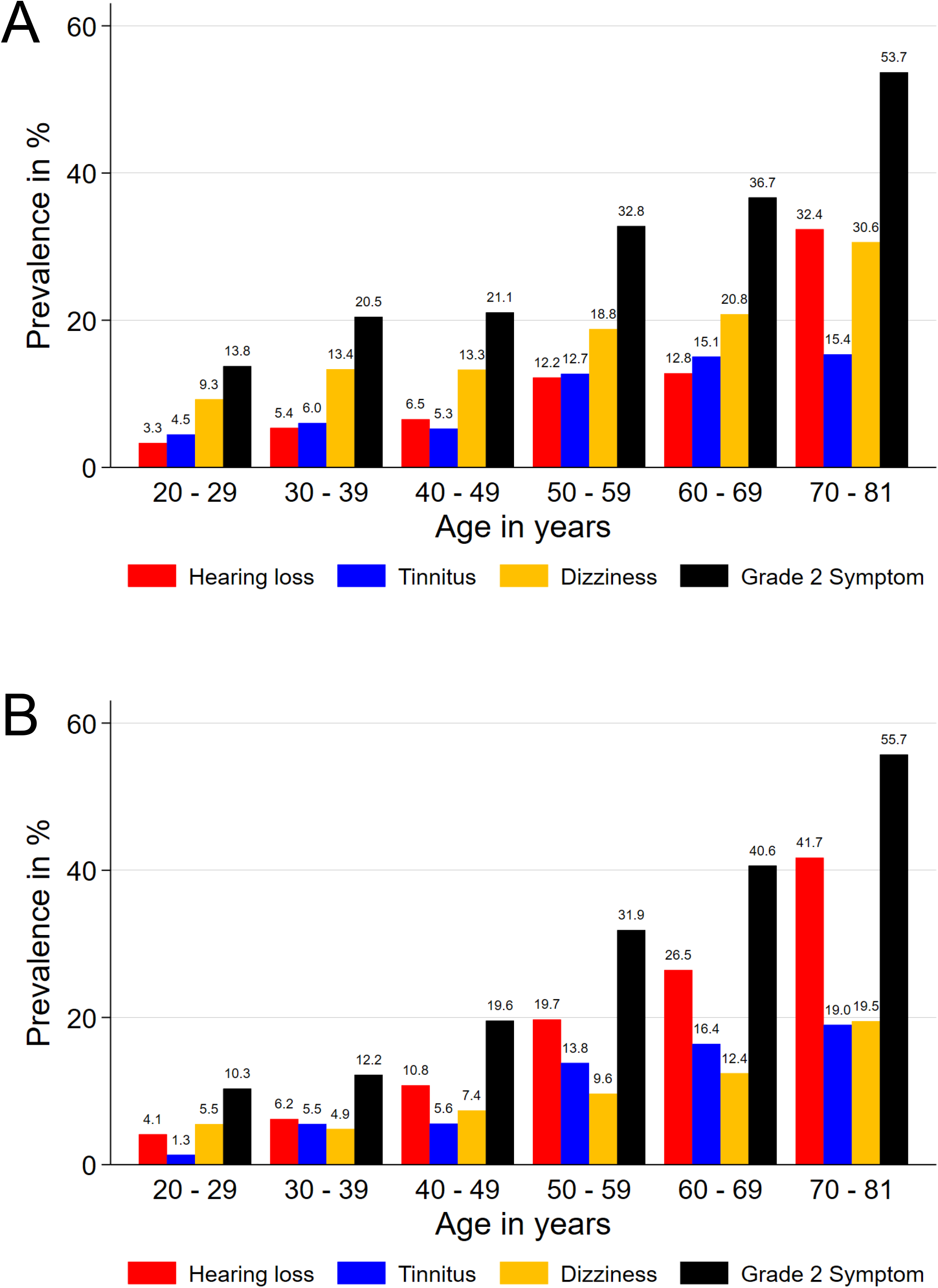
Prevalence by age group and sex. A: female, B: male.

Following this initial exploration, further analyses were adjusted for age and sex (Table 2). Education was the most prominent factor, reducing the odds of any relevant symptoms significantly by 45% when more than 10 years of schooling were compared to less than 10 years. This effect was most pronounced for dizziness, followed by hearing loss. Smoking was the only factor that was significantly associated with all three audiovestibular symptoms as well as the only risk factor significantly associated with tinnitus at all. Of note, this was most distinct when former smokers were compared to individuals that never smoked. The presence of type 2 diabetes as well as higher triglycerides did significantly affect the occurrence of both hearing loss and dizziness. Levels of serum glucose and arterial hypertension significantly increased the odds for hearing loss only. The HbA1c, serum HDL-cholesterol and a diagnosis of dyslipidemia were positively associated with dizziness only.

**Table 2:** Association of behavioural and metabolic markers with audiovestibular symptoms.

|  | **Hearing loss**<br>Odds ratio (95%-CI) | **Tinnitus**<br>Odds ratio (95%-CI) | **Dizziness**<br>Odds ratio (95%-CI) | **Grade 2 symptom**<br>Odds ratio (95%-CI) |
| --- | --- | --- | --- | --- |
| **Education (Years of schooling)** |  |  |  |  |
| **10 years vs. <10 years** | 0.85 (0.71; 1.03) | 0.93 (0.75; 1.15) | 0.76 (0.63; 0.92)* | 0.82 (0.71; 0.95)* |
| **>10years vs. <10 years** | 0.57 (0.46; 0.72)* | 0.79 (0.61; 1.01) | 0.48 (0.38; 0.62)* | 0.55 (0.46; 0.65)* |
| **Smoking status** |  |  |  |  |
| **Former vs. never** | 1.27 (1.06; 1.52)* | 1.24 (1.01; 1.53)* | 1.24 (1.04; 1.48)* | 1.30 (1.13; 1.48)* |
| **Current vs. never** | 1.22 (0.99; 1.50) | 1.09 (0.87; 1.37) | 1.05 (0.87; 1.29) | 1.19 (1.03; 1.38)* |
| **Alcohol consumption; g/day** | 1.06 (0.98; 1.14) | 1.03 (0.95; 1.12) | 0.89 (0.78; 1.01) | 1.01 (0.95; 1.08) |
| **Body mass index; kg/m<sup>2</sup>** | 1.03 (0.96; 1.11) | 1.00 (0.92; 1.09) | 1.04 (0.97; 1.12) | 1.00 (0.94; 1.06) |
| **Waist circumference; cm** | 1.08 (0.99; 1.17) | 1.02 (0.92; 1.12) | 1.07 (0.98; 1.17) | 1.02 (0.96; 1.10) |
| **Glucose; mmol/L** | 1.07 (1.00; 1.14)* | 1.05 (0.96; 1.14) | 1.05 (0.97; 1.13) | 1.05 (0.99; 1.11) |
| **HbA1c; %** | 1.07 (0.99; 1.15) | 1.03 (0.94; 1.13) | 1.10 (1.02; 1.18)* | 1.08 (1.02; 1.15)* |
| **Type 2 diabetes** | 1.37 (1.10; 1.72)* | 1.14 (0.86; 1.49) | 1.48 (1.16; 1.88)* | 1.35 (1.12; 1.64)* |
| **Total cholesterol; mmol/L** | 0.98 (0.90; 1.06) | 1.03 (0.95; 1.13) | 0.94 (0.86; 1.02) | 0.96 (0.90; 1.01) |
| **HDL-cholesterol; mmol/L** | 0.98 (0.89; 1.07) | 0.97 (0.89; 1.07) | 0.91 (0.84; 0.99)* | 0.97 (0.91; 1.04) |
| **LDL-cholesterol; mmol/L** | 0.95 (0.88; 1.03) | 1.04 (0.95; 1.13) | 0.95 (0.88; 1.03) | 0.94 (0.88; 0.99)* |
| **Triglycerides; mmol/L** | 1.11 (1.02; 1.20)* | 1.03 (0.95; 1.11) | 1.09 (1.01; 1.18)* | 1.09 (1.02; 1.16)* |
| **Dyslipidemia** | 1.17 (0.99; 1.39) | 1.22 (0.99; 1.49) | 1.46 (1.22; 1.75)* | 1.33 (1.16; 1.53)* |
| **Arterial hypertension** | 1.23 (1.04; 1.45)* | 1.04 (0.86; 1.25) | 1.12 (0.94; 1.32) | 1.13 (0.99; 1.28) |

2 Data are derived from logistic regression models adjusted for age and gender. Continuous exposure were standardized. CI: confidence interval.
\*: p<0.05.

Subsequently, the factors education and smoking were explored further regarding their relevance for individual and any grade 2 symptoms broken down by sex (Figure 3). While high education reduced the odds for hearing loss, dizziness, tinnitus and any grade 2 symptom in both sexes, medium compared to low education did reduce the odds for all individual as well as any grade 2 symptoms significantly in females only. Former female smokers showed significantly increased odds for hearing loss, tinnitus and any grade 2 symptom in comparison to never smokers, while in males this was the case only for dizziness. There were no significant findings regarding current versus never smokers in both sexes. The most pronounced finding were odds ratios for high versus low education for dizziness in females and hearing loss in males of 0.45 (95% CI 0.30; 0.56) and 0.51 (95% CI 0.39; 0.68) as well as any grade 2 symptoms of 0.51 (95% CI 0.39; 0.66) in females and 0.53 (95% CI 0.42; 0.68) in males, respectively.

**Figure 3:**
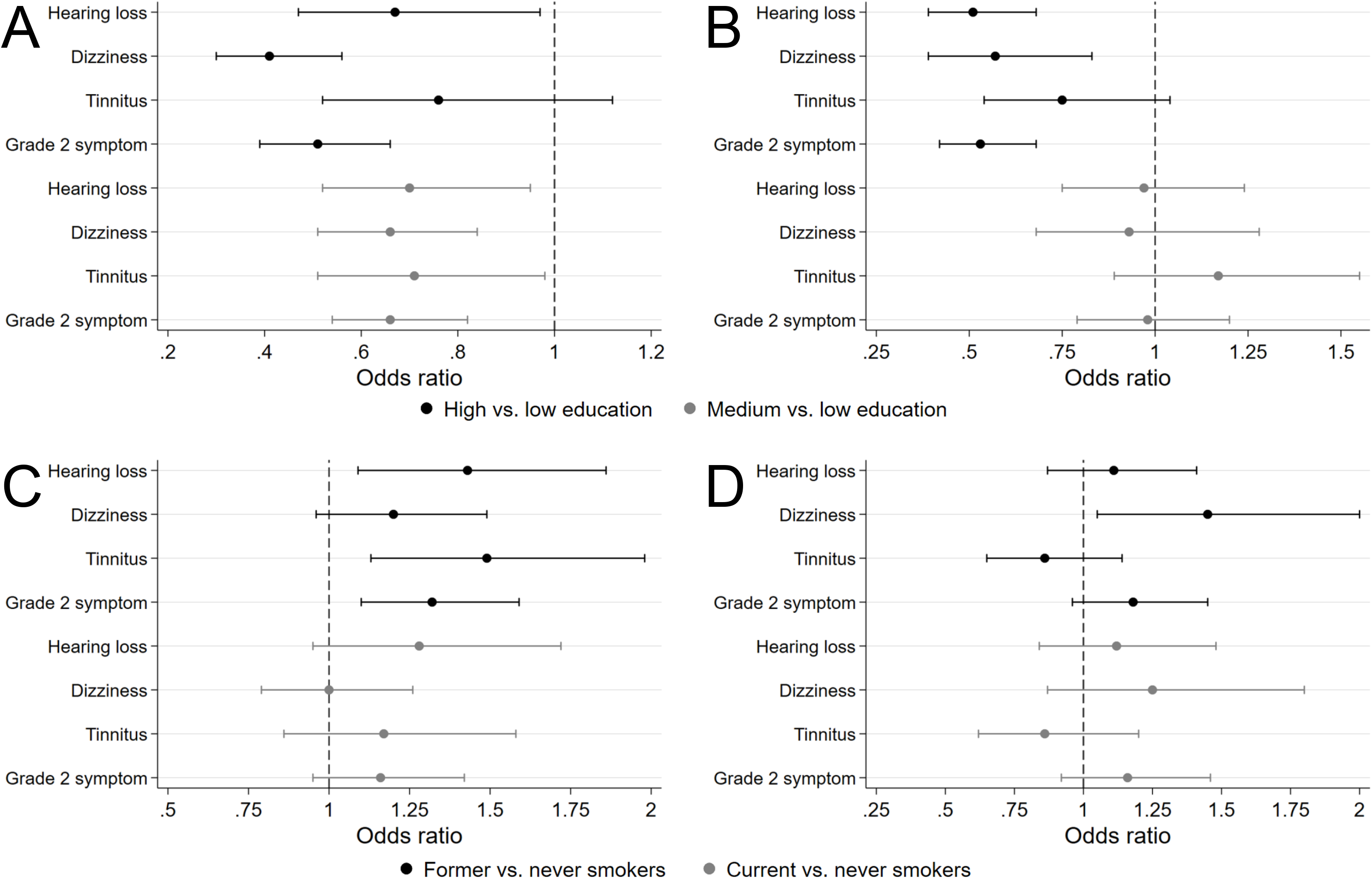
Age-adjusted associations of education and smoking status with audiovestibular symptoms stratified by sex. A, C female, B, D male; A, B education, C, D smoking. Odds ratio (95%-CI).

## Discussion

In our analysis we observed that audiovestibular symptoms were highly prevalent in the investigated cohorts that were drawn from the general population of West Pomerania. A relevant subgroup of affected individuals reported more than one symptom with a moderate correlation between hearing loss and tinnitus. Sex and age had a major influence on the expression of audiovestibular symptoms. Among the investigated individuals, hearing loss was more prevalent in males and dizziness in females. Beyond this, the prevalence rose markedly with advancing age, leading to the majority of individuals at 70 years or older reporting audiovestibular symptoms. The dominant contributor here was hearing loss in both sexes, closely followed by dizziness in females. The prevalence of hearing loss and tinnitus increased significantly over a period of approximately 10 years.

We also studied the influence of several risk factors. Thereby, we found that smoking was associated with the occurrence of all individual audiovestibular symptoms. Less than 10 years of education, a diagnosis of type 2 diabetes and increased triglyceride levels were associated with both hearing loss and dizziness. The risk for hearing loss alone was increased by higher levels of glucose and arterial hypertension while dizziness alone was facilitated by dyslipidemia as well as increased levels of HbA1c. Our results established the prevalence and co-prevalence of audiovestibular symptoms in a population-based sample. It is important to note that each symptom has a different set of possible causes with a considerable overlap between symptoms. Additionally, a wide range of topographic locations of the ear or associated systems may be involved.

Regarding diagnostic assessment, fundamental differences exist between the audiovestibular symptoms studied here. For tinnitus, a subjective sensation of the auditory pathway by definition, no objective diagnostic test exists (Jackson et al. 2019). Thereby, the findings presented here are a best possible estimation of the prevalence and severity of tinnitus in the population. Furthermore, self-report is an established measure for tinnitus in epidemiological studies (McCormack et al. 2016). Contrastingly, self-reported hearing loss and dizziness or vertigo must not be confused with middle or inner ear dysfunction as determined by audiological or vestibular diagnostic tests. Self-reporting is, however, established for the assessment in clinical and research contexts for hearing loss as well as for dizziness.

In screening for hearing loss, the value of a single question is well established and reaches a sensitivity and specificity of 80% and 74%, respectively (Feltner et al. 2021). Likewise, in epidemiological studies, self-reported hearing loss was validated as a reasonable estimate for audiologically determined hearing loss as standard mode of assessment: In a brief review accompanying the reporting of an original investigation of hearing loss in an older population, Sindhusake and coworkers summarized for different short questions on subjective hearing loss a sensitivity of 66 – 78% and a specificity of 67 – 80% for audiometrically proven mild hearing loss. For moderate hearing loss, the sensitivity was given as 90 – 93% and the specificity as 56 – 71% (Sindhusake et al. 2001). This association, however, has found to be frequency-dependent (Hannula et al. 2011), and may be further explained by age and sex (Kamil et al. 2015). Further discrepancies as over- or underreporting of audiometrically determined hearing loss result from exposure to occupational noise, hypotension, and depression (Choi et al. 2019). In a study with very detailed characterization of hearing, 20.2% of the subjects with self-reported hearing loss did not even meet the definition of mild hearing loss in the pure-tone audiogram while 6.2% of the individuals reporting no hearing problem did have at least mild hearing loss in audiometry (Gates et al. 1990). It seems, however, possible that self-reported hearing loss is sensitive for hearing impairment that does not yet yield pathological results in pure-tone audiometry or other diagnostic tests (Kamerer et al. 2022). This suggests that self-reported hearing represents a different spectrum of auditory function and potentially central processing in contrast to pure-tone audiometry.

For dizziness or vertigo, simple or structured questions are considered to have unsatisfactory sensitivity and specificity for pathological findings in subsequent vestibular testing. Pathological responses in a widely used questionnaire for vestibular problems in epidemiological research have been found to predict abnormal vestibular test results rather poorly (Cohen et al. 2022). A differentiation of peripheral vestibular dysfunction from central, functional or psychiatric disorders, however, was out of the scope of this work. Neuhauser and coworkers identified a lifetime prevalence of 29.5% for moderate or severe vertigo or dizziness in the population by a single question that was comparable to our approach. Of note, in a focused neurotological interview thereafter, 24.2% of those individuals reported symptoms that were suggestive of a peripheral vestibular disorder (Neuhauser et al. 2005). It is likely that a comparable distribution underlies the dizziness complaints determined in our population. However, to clarify this, either a specialized interview or, preferably, vestibular diagnostic tests have to be performed. Regarding self-report, the exact phrasing of a question on balance disorders is complex and carries cultural as well as linguistic implications. This has been discussed previously in particular for the difference between the words ‘dizziness’ and ‘vertigo’, assuming that the latter might be more specific for a rotational perception and thereby for peripheral vestibular disorders (Murdin & Schilder 2015). Despite considerable progress in this question by consented definitions (Bisdorff et al. 2009), there is still no universally accepted solution that would be viable for epidemiological investigations in different language areas. While acknowledging the limitations of self-reported symptoms in the absence of diagnostic tests, they do capture the whole and thereby a more complex processing pathway from basic sensory perception to cognition, including emotional assessment and mood.

In our general population sample, we observed a weighted prevalence of 14.2% for hearing loss, 9.7% for tinnitus and 13.5% for dizziness that is generally in line with previous reports. Inherently to the nature of tinnitus, an assessment by a single question is the standard method in clinical and epidemiological studies and usually represents a point prevalence. This results in prevalence ranges reported from 5.1 to 42.7% when asked without qualifier and 3.0 to 30.9% when only bothersome tinnitus is considered (McCormack et al. 2016). A systematic review and meta-analysis resulted in a pooled prevalence of 14.4% (95% CI 12.6; 16.5) for any tinnitus and 2.3% (95% CI 1.7; 3.1) for severe tinnitus (Jarach et al. 2022).

Comparable to the methodical considerations given above, the situation for published prevalence estimates is different for hearing loss and dizziness. In population-based studies that included all age ranges of adult individuals, the prevalence of self-reported hearing loss ranged from 13.0 to 26.8% in studies from Eastern Asia and Northern America (Choi et al. 2019; Dillard et al. 2022), placing our result within this range of developed countries with high life expectancy. This is, however, considerably higher than a global estimate that places the prevalence of moderate to severe hearing loss at 5.2% globally and 5.8% for the European region, where the present study originated. Those notably excluded self-reported hearing loss and thereby only considered prevalence figures based on audiological assessment (Orji et al. 2020). Another study extrapolated a prevalence of 16.2% for mild and 5.8% for moderate hearing loss for Germany based on two different investigations in three samples based on audiometry (Gablenz et al. 2017).

The estimates for the prevalence of dizziness without consideration of a potential peripheral vestibular origin range from 15 to 35% (Murdin & Schilder 2015; Maudoux et al. 2022). When vertigo is considered separately and more specific as dysfunction of the peripheral vestibular organ in the inner ear, 3 - 10% are given (Murdin & Schilder 2015). Neuhauser and coworkers identified a lifetime prevalence of 29.5% for moderate or severe vertigo or dizziness in the German population by a single question that was comparable to our approach. By employing those terms in a community-based study from north-eastern France, the view of “vertigo” as a term for primarily vestibular dysfunction as a subset of balance dysfunction is even more challenged by a reported one-year prevalence of vertigo of as high as 48.3%, while unsteadiness and dizziness were noted by only 39.1% and 35.6% of individuals, respectively (Bisdorff et al. 2013).

While it is consistently assumed that the audiovestibular symptoms hearing loss, tinnitus and dizziness are very common in the population, the wide variance among prevalence numbers given in the literature is explained by differences in the definition of disorders and cut-off values in diagnostic tests as well as different methods of assessment (Murdin & Schilder 2015; McCormack et al. 2016; Sheffield & Smith 2019). Additionally, the prevalence can be assumed to be heavily influenced by factors like age, as determined by the age distribution in the underlying population. Likewise, studies on older populations typically yield a higher prevalence of symptoms. Furthermore, point prevalence, period prevalence and lifetime prevalence are often not clearly distinguishable by self-report. An analysis from the Framingham cohort with 1662 individuals with an age beginning in the 6^th^ decade of life and older described self-reported hearing loss in 41.1%, audiometrically determined mild hearing loss in 29% and tinnitus in 16.8% (Gates et al. 1990). The Blue Mountains Hearing Study, that included individuals from the age of 49 years and older, reported a prevalence of 39.7%, 13.9% and 2.3% for mild, moderate and severe self-reported hearing loss, respectively (Sindhusake et al. 2001).

We found an increase in the prevalence of hearing loss and tinnitus in both sexes as well as in dizziness in men between the two cohorts of SHIP that were assessed in this study. Changing prevalence within the comparable short timeframe of about 10 years that separates those cohorts has been reported earlier for thyroid disorders (Khattak et al. 2016) as well as lifestyle-related risk factors (Völzke et al. 2015). Those trends could be related to social development, health-related legislation and public-health programs. Contrastingly, increasing education as well as reduced occupational noise exposure, ear infections and smoking in the population are considered to lead to less ear disorders (Engdahl, Stigum, & Aarhus 2021). A likewise observation has been made by an actual decrease of age- and sex-specific hearing loss in a comparison of pure-tone audiometry results between two cohorts investigated 20 years apart in Norway (Engdahl, Strand, & Aarhus 2021). However, for self-reported measures increased awareness or higher health expectations in more recent generations may also play a role. That has been brought forward as an explanation of an age-specific increase in the prevalence of tinnitus in age groups that were five years apart despite an overall declining prevalence of hearing loss in a study from Wisconsin, USA (Nondahl et al. 2012).

Education was the most prominent factor influencing the occurrence of audiovestibular symptoms in our study. The odds of any relevant symptom as well as hearing loss and dizziness individually were significantly reduced when more than 10 years of schooling were compared to less than 10 years. This may be a sign for a shared pathway of neurodegeneration affecting cognition as well as sensory systems (Brenowitz et al. 2019). The auditory pathway might be particularly susceptible (Uchida et al. 2019; Johnson et al. 2021), but anatomical and functional similarities exist between age-related changes in the cochlear and vestibular part of the inner ear (Paplou et al. 2021). Cognitive stimulation, as approximated by education level reached during life and strongly influenced by hearing and communication abilities, is a major factor influencing dementia risk in later life (Livingston et al. 2020). Neurodegeneration in turn may relate to hearing loss bi-directionally not only by sensory deprivation, but also by simultaneous degenerative processes at multiple stages of the auditory processing pathway (Johnson et al. 2021).

In our study, several cardiovascular risk factors were investigated as an association to hearing or balance dysfunction was assumed due to numerous references in the literature. Thereby, associations with smoking, diabetes, dyslipidemia and hypertension were found. Smoking was the only single factor that was significantly associated with all three audiovestibular symptoms. While in the past an association with smoking has been reported mostly individually for hearing loss (Cruickshanks et al. 1998; Lin et al. 2013; Tsimpida et al. 2019; Engdahl, Stigum, & Aarhus 2021; Tseng et al. 2022; Mick et al. 2023), tinnitus (Biswas et al. 2021; Biswas & Hall 2021; Goderie et al. 2022) and dizziness (Neuhauser et al. 2005; Agrawal et al. 2009), the underlying cause has not been clarified so far. In an animal model of oxidative stress, degeneration and loss of cochlear spiral ganglion neurons have been documented after chronic exposure to cigarette smoke (Paquette et al. 2018). This mechanism might be equally relevant for the development of tinnitus or dizziness, as other neuronal tissues beyond the cochlea and auditory nerve may also become degraded.

In previous reports, diabetes mellitus was associated with dysfunction of the auditory and vestibular systems (Agrawal et al. 2009; Horikawa et al. 2013; Biswas & Hall 2021; Samocha-Bonet et al. 2021; Tseng et al. 2022). It has been described in relation to hearing loss (Horikawa et al. 2013; Samocha-Bonet et al. 2021; Tseng et al. 2022; Mick et al. 2023), balance disturbance (Agrawal et al. 2009) as well as tinnitus (Biswas & Hall 2021). In our data, however, diabetes was linked to hearing loss and dizziness but not to tinnitus. A similarly missing link between tinnitus and diabetes mellitus has been attributed to a lack of power by low exposure in a previous study (Goderie et al. 2022). The complex relationship between diabetes mellitus and hearing loss has been explored in particular detail in the past and likely includes a multitude of different routes like toxic effects of hyperglycemia, diabetic microangiopathy and neuropathy as well as side-effects of diabetic medications (Samocha-Bonet et al. 2021). Most likely, a diabetes-associated affection of hearing takes already place at a precursor stage of diabetes (Seo et al. 2016). For the relationship between diabetes and balance dysfunction, a possible role for central insulin resistance has been suggested (Case et al. 2022).

The association found here between dyslipidemia as well as hearing loss and dizziness confirms earlier reports in this regard (Neuhauser et al. 2005; Nash et al. 2011), but a detailed mechanism is still warranted. In a mouse model of hyperlipidemia, the development of hearing loss could be prevented by a statin while simultaneously oxidative stress and cell death in the cochlea were reduced (Lee et al. 2020). A cross-sectional study that explicitly addressed dyslipidemia did not find it to be directly associated with hearing loss. However as one of three cardiovascular risk factors, the others being hypertension and diabetes, it did increase the prevalence of hearing impairment significantly when two or all of the three factors were present (Hara et al. 2020).

A diagnosis of arterial hypertension was only significantly associated with hearing loss in our data, confirming earlier studies (Tseng et al. 2022; Mick et al. 2023). Previous reports linked this condition to tinnitus (Biswas & Hall 2021) and dizziness (Neuhauser et al. 2005; Agrawal et al. 2009; Nash et al. 2011) as well. Looking at our findings on cardiovascular risk factors and audiovestibular dysfunction in the light of the body of evidence from the literature, a strong but complex link can be assumed. Due to a multifaceted interaction, the effect sizes may be too small to be assessed by surrogate markers in individual studies.

An interaction between function and dysfunction of the senses of hearing and balance is widely assumed. By calculating the phi-coefficient, we encountered a moderate positive relationship between hearing loss and tinnitus with inconclusive results for the other two pairings. Contrastingly, a population-based cross-sectional study in individuals of 50 years and older found a significant association between dizziness and tinnitus, but not between dizziness and hearing loss (Gopinath et al. 2009). In a clinical setting, age-related high frequency-hearing loss was not associated to measurable peripheral vestibular dysfunction (Schubert et al. 2022), emphasizing that the detailed mechanism of interaction between auditory and vestibular dysfunction is still elusive. It is, however, necessary to gain more insight in the future, since multisensory impairment is reported to be strongly associated with neurodegeneration, an association that has been already found for vision, hearing, smell, and touch (Brenowitz et al. 2019) and could well be applicable to the interaction of hearing and balance also.

## Conclusion

In a population-based study of adults, self-reported hearing loss, tinnitus and dizziness were highly prevalent. A moderate interaction was found between hearing loss and tinnitus. Age and sex were major contributing factors. In adjusted regression analyses, smoking increased the odds ratio for all three symptoms. Less education as well as different cardiovascular risk factors contributed to hearing loss and dizziness. Further studies are warranted to clarify the complex interaction between those three audiovestibular symptoms as well as more details on the mechanism of influencing factors.

## Data Availability

All data produced in the present study are available upon reasonable request to the authors.

## Acknowledgements

The authors are grateful to the participating population of Western Pomerania and to the team involved in the conception and conduction of SHIP-START and SHIP-TREND.

The Study of Health in Pomerania (SHIP) is part of the Community Medicine Research Network of the University of Greifswald, which is supported by the Ministry of Cultural Affairs as well as the Social Ministry of the German Federal State of Mecklenburg-West Pomerania. SHIP was also funded by the German Federal Ministry of Education and Research (BMBF) with the grant numbers: 01 ZZ 9603, 01 ZZ 0103, 01 ZZ 0403, and 01 ZZ 0701.

Parts of this work were presented at the 94th Annual Meeting of the German Society of Oto-Rhino-Laryngology, Head and Neck Surgery, May 17 to May 20, 2023, in Leipzig, Germany (https://doi.org/10.1055/s-0043-1767437).

## Supplement

<u>Supplemental Table 1: Completed checklist of items to be included for cross-sectional studies according to the STROBE Statement</u>

<u>Supplemental Table 2: Phrasing and grading of primary outcome measures</u>

<u>Supplemental Table 3: Excluded participants and analyzed population</u>

