## Supplemental Table 2 for "Prevalence and risk factors of self-reported audiovestibular symptoms in a population-based sample from rural northeastern Germany"

**Supplemental Table 2: Phrasing and grading of primary outcome measures**

| | Questions | Options | | | |
|---|---|---|---|---|---|
| Introductory remarks for complaints that include hearing loss and dizziness.<br>*(English translation)* | Im Folgenden wird eine Reihe von Beschwerden genannt. Machen Sie bitte bei jeder aufgeführten Beschwerde ein Kreuz in eines der vier Kästchen, je nachdem, ob sie gar nicht, kaum, mäßig oder stark unter diesen Beschwerden leiden.<br>*In the following several of disorders will be named. Please give an answer to every below-mentioned disorder and decide where to tick depending on how strong you suffer from these symptoms.* | | | | |
| Hearing loss<br>*(English translation)* | Schwerhörigkeit, Hörbeschwerden<br>*Hearing loss, hearing difficulties* | Gar nicht<br>*Not at all* | Kaum<br>*Rarely* | **Mäßig**<br>***Moderately*** | **Stark**<br>***Severely*** |
| Dizziness<br>*(English translation)* | Schwindelgefühl<br>*Dizziness* | Gar nicht<br>*Not at all* | Kaum<br>*Rarely* | **Mäßig**<br>***Moderately*** | **Stark**<br>***Severely*** |
| Introductory remarks for complaints that include tinnitus<br>*(English translation)* | Leiden Sie an den im folgenden genannten Beschwerden?<br>*Do you suffer from one of the following complaints?* | | | | |
| Tinnitus<br>*(English translation)* | Ohrgeräusche, Ohrensausen<br>*Tinnitus/ ringing in the ears* | Nein<br>*No* | Manchmal<br>*Sometimes* | **Häufig**<br>***Frequently*** | **Immer**<br>***Always*** |
| Severity grade | | 0 | 1 | **2** | **3** |

The severity grades 2 and 3 (**bold**) were considered as relevant values for the analysis in this study.
