## Supplemental Table 3 for "Prevalence and risk factors of self-reported audiovestibular symptoms in a population-based sample from rural northeastern Germany"

Supplemental Table 3: Excluded participants and analyzed population

| | excluded from analysis (n = 593) | analyzed population (n = 8134) | p* |
|---|---|---|---|
| **Females** | 56.3% | 50.8% | 0.009 |
| **Age; years** | 62 (46; 73) | 51 (38; 63) | <0.001 |
| **Age groups; years** | | | <0.001 |
| **20-30** | 4.9% | 11.1% | |
| **30-40** | 13.5% | 17.0% | |
| **40-50** | 11.5% | 19.3% | |
| **50-60** | 15.7% | 20.0% | |
| **60-70** | 19.9% | 18.8% | |
| **70-80** | 34.6% | 13.9% | |
| **Education** | | | <0.001 |
| **< 10 years** | 45.2% | 30.6% | |
| **= 10 years** | 42.8% | 47.9% | |
| **> 10 years** | 12.0% | 21.5% | |
| **Smoking** | | | 0.155 |
| **Current smoker** | 38.1% | 36.0% | |
| **Former smoker** | 36.9% | 35.2% | |
| **Never smoker** | 25.0% | 28.8% | |
| **Alcohol consumption; g/day** | 1.5 (0.0; 6.5) | 4.6 (1.1; 13.1) | <0.001 |
| **Body mass index; kg/m²** | 28.6 (25.3; 32.3) | 27.1 (24.0; 30.5) | <0.001 |
| **Waist circumference; cm** | 94 (84; 105) | 90 (79; 100) | <0.001 |
| **Diabetes mellitus** | 16.1% | 8.8% | <0.001 |
| **Hypertension** | 65.6% | 49.2% | <0.001 |
| **Dyslipidemia** | 25.7% | 21.4% | 0.016 |

Data are expressed as median, $25^{th}$ percentile, $75^{th}$ percentile (continuous data) or as percentage (categorical data)

* Mann-Whitney-U test (continuous data) or $\chi 2$-test (categorical data)
